# The relationship of glutamate and glutamine and metabolic profiling in focal epilepsy using 7T CRT-FID-MRSI

**DOI:** 10.1101/2025.02.12.25321529

**Authors:** Stefanie Chambers, Philipp Lazen, Haniye Shayeste, Matthias Tomschik, Jonathan Wais, Lukas Hingerl, Bernhard Strasser, Lukas Haider, Tatjana Traub-Weidinger, Christoph Baumgartner, Johannes Koren, Florian Mayer, Martha Feucht, Christian Dorfer, Ekaterina Pataraia, Wolfgang Bogner, Siegfried Trattnig, Gregor Kasprian, Karl Rössler, Gilbert Hangel

**Affiliations:** Department of Neurosurgery, Medical University of Vienna, Austria; MRCE, Department of Biomedical Imaging and Image-guided Therapy, Medical University of Vienna, Austria; Christian Doppler Laboratory for MR Imaging Biomarkers, Vienna; Division of Neuroradiology and Musculoskeletal Radiology, Department of Biomedical Imaging and Image-guided Therapy, Medical University of Vienna, Austria; NMR Research Unit, Queen Square Multiple Sclerosis Centre, Queen Square Institute of Neurology, University College London, United Kingdom; Division of Nuclear Medicine, Department of Biomedical Imaging and Image-guided Therapy, Medical University of Vienna, Austria; Department of Neurology, Klinik Hietzing, Vienna, Austria; Department of Pediatrics and Adolescent Medicine, Medical University of Vienna, Austria. Full Member of ERN EpiCARE; Department of Neurology, Medical University of Vienna, Austria; Functional Imaging Laboratory, UCL Queen Square, Institute of Neurology, University College London, United Kingdom

**Keywords:** Epilepsy, ultra-high-field MRSI, spectroscopy, 7T

## Abstract

Approximately one-third of people with epilepsy (PWE) remain drug-resistant. In these cases, surgical resection of the epileptogenic zone may significantly reduce or eliminate seizures. Surgery necessitates precise delineation of the epileptogenic zone which proves especially challenging in the 20% of PWE that remain MRI-negative. To this end, the purpose of this study was to analyze the feasibility and robustness of ultra-high-field MRSI in identifying and characterizing pathologies in focal epilepsy. In addition, the relationship of glutamate and glutamine was evaluated in the epileptogenic zone (EZ)

Fifty-six people with focal epilepsy were prospectively measured using 7T concentric ring trajectory direct acquisition of free-induction-decay MRSI which generated whole-brain metabolic maps with an isotropic resolution of 3.4mm^3^. After exclusion criteria were applied, we assessed metabolite ratios in 29 lesional- and MRI-negative PWE.

In the lesional group, metabolic alterations in the suspected EZ were present in 86.7% of maps normalized to NAA, whereas this was reduced to 80% in creatine ratios. Metabolites with the highest stability in the lesional group included myo-inositol and choline, increased in 92.3%. In MRI-negative patients, changes were heterogeneous and less circumscribed, with a detection rate of 57.1%. We also observed a tendency towards an inverse relationship of glutamate to glutamine in the EZ, with relative increases of glutamine in PWE with lower seizure frequencies, contrasting relative glutamate increases in higher seizure frequencies.

Our preliminary analysis suggests that 7T CRT-FID MRSI shows promise not only in identifying metabolic alterations in focal epilepsy but may also provide insights into disease pathomechanisms.

## 1. Introduction

Epilepsy is caused by a wide array of underlying pathologies and approximately one-third of people with epilepsy (PWE) remain drug-resistant [1] despite the use of multiple antiseizure medications (ASM). In people with focal epilepsy (PWFE), surgery may prove beneficial, but post-operative seizure freedom heavily depends on total resection of the epileptogenic zone [2]. In some cases this zone may extend past visible lesion borders or remain undetectable in structural MRI (Magnetic Resonance Imaging), referred to as MRI-negative focal epilepsy. Though a recent implementation of the 7T MRI consensus protocol [3] for epilepsy showed that the application of ultra-high-field strengths yields a diagnostic gain of up to 50% in focal epilepsy compared to dedicated 3T epilepsy protocols [4], the remaining medically non-responsive patients without macroscopically detectable lesions pose a challenge in surgical planning. In such cases, imaging modalities that shed light on brain functionality by measuring metabolites [5] (such as positron-emission tomography (PET) or magnetic resonance spectroscopy (MRS)) may aid in identifying subtle pathological alterations that are otherwise blind to structural MRI. However, given that placement of the ROI is necessary due to the circumscribed coverage of MRS, its practicality in MRI-negative cases in which the epileptogenic zone (EZ) is frequently unclear is limited. As these patients make up 20% of the cohort [6], there is a pressing need for the application of whole-brain metabolic maps with larger volume coverage while maintaining spectral resolution.

In addition, MRS in epilepsy in routine clinical use has frequently been limited by overlapping resonance frequencies of metabolites due to low spatial resolution and signal-to-noise ratios (SNR) at standard clinical field strengths (i.e. 1.5T and 3T), [7], [8]. Ultra-high field (UHF) MR systems, such as 7T, can overcome some of these limitations through increased spatial resolution due to higher SNR and the separation of metabolites with otherwise overlapping resonance frequencies such as glutamate (Glu) or glutamine (Gln) (otherwise subsumed as Glx at lower field strengths). However, higher field strengths incur their own set of problems, such as increased B_o_ and B_1_-inhomogeneities, fast 2T*-relaxation times and higher specific absorption times, limiting their applicability in routine clinical practice.

To this end, we have developed a fast, high-resolution MRSI method based on concentric ring trajectory direct acquisition of free-induction-decay (CRT-FID) [9], [10], [11], [12], which enables the generation of 3D whole-brain maps of 14 neurochemicals at 3.4 mm isotropic resolution within 15 minutes, making high-resolution whole-brain MRSI feasible in PWFE for the first time. Thus far, we have successfully applied this method to neurological diseases such as multiple sclerosis [13], [14] and brain tumors [15], [16].

With respects to epilepsy, MRS research to date has largely been limited by the use of single-voxel and multi-slice spectroscopy [7], [8] which is useful in patients with visible MRI lesions and in whom ROI placement is evident. MRI-negative cases, however require whole-brain coverage and, although this has been achieved at lower field strengths (3T) [17], it is limited by decreased spatial resolution and overlapping resonance frequencies of key metabolites in epilepsy, such as Glu and Gln. Because epilepsy is a highly metabolic illness with alterations potentially representing both cause and consequence of hyperexcitability [18], a body of research has been dedicated to deciphering such changes. Research in focal epilepsy has largely focused on metabolic changes in hippocampal sclerosis and malformations of cortical development (MCD) and in this most prominently, focal cortical dysplasia (FCD). Groups investigating metabolic alterations using MRS in MCD [8], [19] have found N-acetyl-aspartate (NAA), synthesized in neuronal mitochondria and a measure of healthy neuronal function [20], [21] to be significantly decreased in FCD, a phenomenon that has been described over numerous focal epileptic lesions [8], [17], [20], [21]. Decreases in NAA are often mirrored by increases in choline (Cho) [19], a marker for cell membrane turnover, frequently altered in tumors, and myoinositol (mIns) [8], reflecting astroglial hypertrophy. However, the metabolic landscape throughout the EZ remains ambiguous, highlighting the complexity of the dynamic processes inherent to epileptic metabolic activity as well as its numerous drivers. Some metabolites have been shown to be altered to various extents dependent on clinical parameters, such as the seizure frequency, as seen in creatine [8], an energy buffer, for which increases [8], [22] or decreases [19] have been described. Another key metabolite and neurotransmitter with diverse functions in neurotransmission and energy metabolism that is inherently dependent on clinical variables is Glu, closely linked to Gln through the glutamate-glutamine cycle. Increases [23], unchanged concentrations [24], as well as decreases for Glx have been described interictally, though previous in-vivo research has frequently been limited by the overlapping resonance frequencies of these neurochemicals, therefore compounding them as the entity “Glx”. However, these metabolites may act in a countervailing manner in energy metabolism [23], [25], therefore necessitating their spectral separation for adequate interpretation. This is supported by the findings of the application of 7T CRT-FID MRSI in tumors, showing decreases of peritumoral Glu/Gln ratios in tumor-associated epilepsy (TAE), in contrast to those without TAE [26]. In addition, Gln has been shown to be increased in tumor tissue, potentially to uphold metabolism in anaerobic conditions, whereas increases in Glu have been linked to tumor proliferation [15], further highlighting the necessity of their spectral separation.

The heterogeneity of the changes described above highlights the need for ultra-high-resolution, whole-brain MRSI, which enables the detection of epileptogenic zones and interpretation of metabolic changes through sufficient spatial resolution. The aim of our study was, therefore, to qualitatively analyze the feasibility and robustness of 7T CRT-FID MRSI at 7T in focal epilepsy to generate high-resolution 3D maps spanning the cerebrum. Its use as a diagnostic, and potentially therapy-monitoring marker will be assessed. Specifically, we investigated its acuity in the identification and characterization of metabolic alterations over pathologies related to focal epilepsy and assessed the relationship of glutamate and glutamine in epileptic foci.

## 2. Methods

### 2.1. Patient selection criteria and clinical work-up

Following the approval of the institutional board of review (EK 1039/2020) 56 PWFE of at least 12 years of age (range, 14-57 years, 26 females/30 males) were enrolled in this prospective study following written, informed consent (or that of their guardian). Exclusion criteria included contraindications for measurements at 7T (such as claustrophobia, metal implants or pregnancy) and vascular etiologies. PWFE who were included in this study had all undergone an extensive presurgical clinical work-up [27], [28] at the Medical University of Vienna or Department of Neurology, Klinik Hietzing, Vienna, including prolonged a video-electroencephalogram(VEEG), assessment of seizure semiology, neuropsychological testing, a 3T MRI using a dedicated epilepsy protocol and [¹⁸F]Fluorodeoxyglucose (FDG)-PET to localize the suspected epileptogenic zone (EZ). For the purpose of this preliminary analysis, the suspected EZ was expanded to the whole affected lobe. All patients had been presented at interdisciplinary board meetings consisting of neuroradiologists, adult and pediatric epileptologists and neurosurgeons for the evaluation of surgical candidacy. All fifty-six patients were prospectively measured from July 2020 to July 2024 prior to surgery and off these, ten were consequently operated on, making histopathological verification of the diagnoses available. We dichotomized the patients into a lesional (either suspected, after clinical neuroradiological assessment, or histopathologically confirmed lesion) and non-lesional (i.e., MRI-negative) group. The clinical parameters evaluated in the course of this study included the seizure frequency, quantified using the seizure frequency score [29] (categorized into daily = 9-10, weekly = 8, monthly = 7 and yearly occurring seizures = 5-6) and epilepsy duration (**Table 1**). A detailed overview of the clinical characteristics of included patients is provided in **Suppl. Table 1**. A flow-chart of patient recruitment is provided in **Figure 1**.

**Fig. 1.**
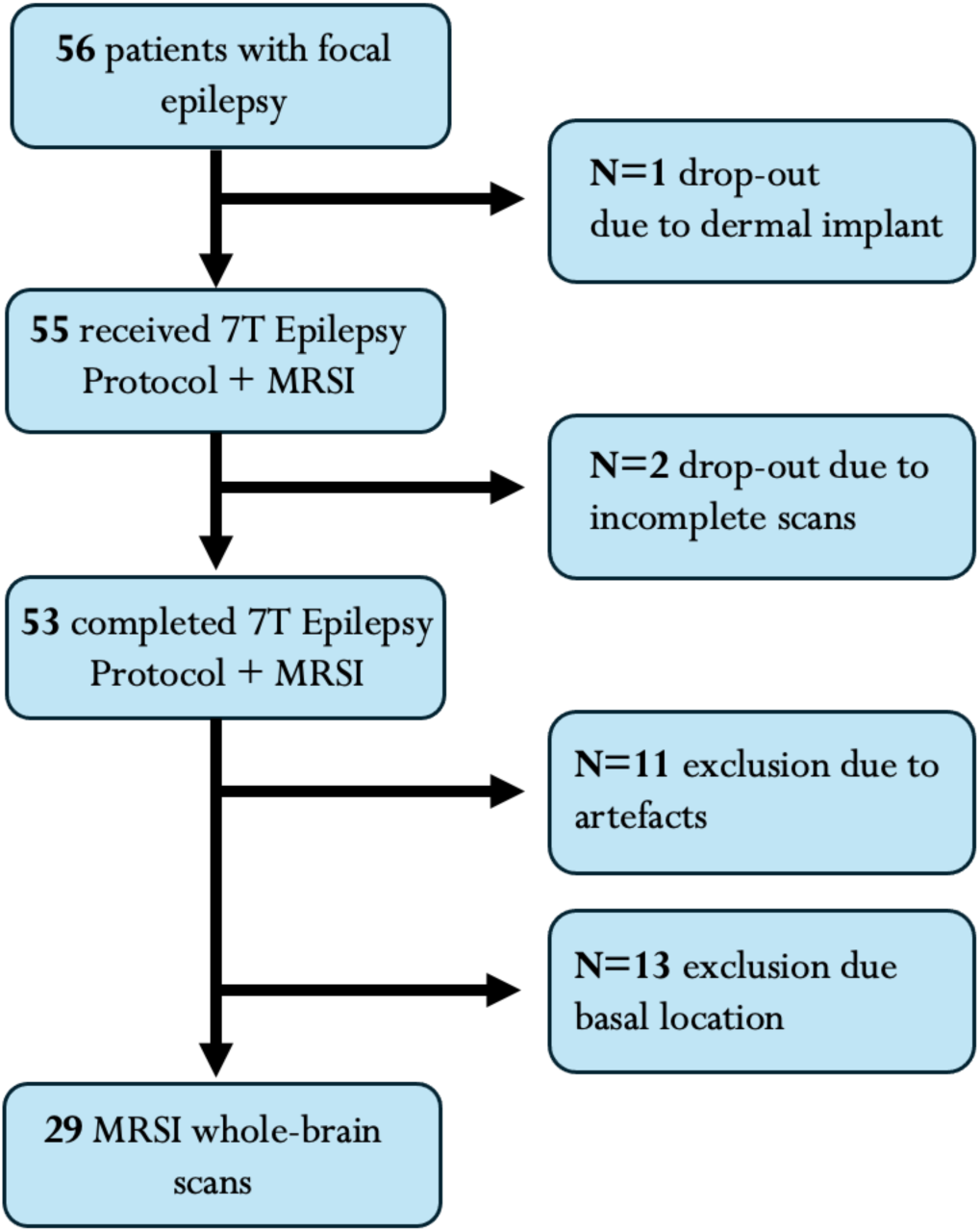
Flow-chart of recruitment and drop-outs during measurement and analysis. Exclusion criteria included the presence of movement or lipid artefacts in derived maps and basally located (temporomesial) EZ, due to impeding field inhomogeneities. <u>Abbreviations:</u> 7T = 7 Tesla, MRSI = Magnetic Resonance Spectroscopy Imaging, EZ = epileptogenic zone

**Table 1:**
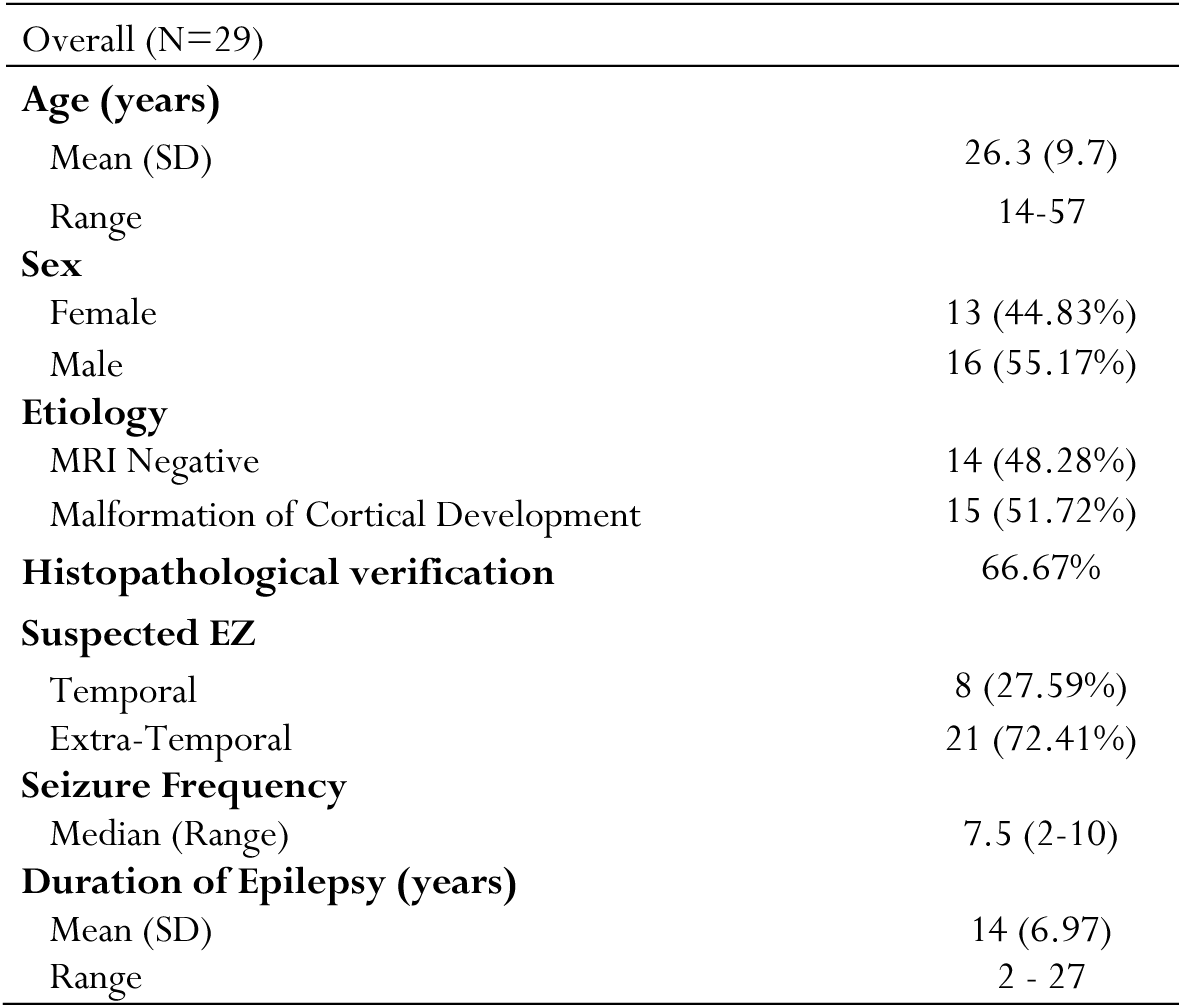
Cohort Summary. **Overview of patient cohort**. MRI-negative excluded incidental findings unrelated to the EZ. An extensive table of included pathologies of Malformations of Cortical Development and affected lobes in extra-temporal epilepsy is provided in **Suppl.** Table 1.

|  |  |
| --- | --- |
| Overall (N=29) |  |
| **Age (years)** |  |
| Mean (SD) | 26.3 (9.7) |
| Range | 14-57 |
| **Sex** |  |
| Female | 13 (44.83%) |
| Male | 16 (55.17%) |
| **Etiology** |  |
| MRI Negative | 14 (48.28%) |
| Malformation of Cortical Development | 15 (51.72%) |
| **Histopathological verification** | 66.67% |
| **Suspected EZ** |  |
| Temporal | 8 (27.59%) |
| Extra-Temporal | 21 (72.41%) |
| **Seizure Frequency** |  |
| Median (Range) | 7.5 (2-10) |
| **Duration of Epilepsy (years)** |  |
| Mean (SD) | 14 (6.97) |
| Range | 2 - 27 |

### 2.2. Measurement protocol

The 3D-MRSI protocol was acquired with a 7T scanner (Siemens Healthineers, Magnetom Plus, Erlangen, Germany) with a 32Rx/1Tx-coil (Nova Medical, Wilmington, MA, USA). The MRSI sequence [11] using 2D-concentric ring trajectories (CRT) featured a 64×64×39 measurement matrix and a 220 × 220 × 133 mm³ field of view (FOV) resulting in 3.4 mm isotropic resolution acquired in 15 minutes. It used free induction decay (FID) acquisition with a 39° flip angle and an acquisition delay of 1.3ms. A TR of 450 ms and WET water suppression [12] allowed for an SNR-optimized readout of 345 ms and a 2778 Hz spectral bandwidth. Details according to Minimum Reporting Standards in in vivo Magnetic Spectroscopy (MRSinMRS) [30] are supplied in Suppl. Table 4. In addition to the MRSI protocol, morphological sequences included MP2RAGE, coronal hippocampal T2-weighted images, 3D FLAIR, 3D WMS, and transversal SWI, as previously published [4].

### 2.3. Data post-processing

Following the acquisition, the MRSI data was processed with an in-house pipeline [31] including lipid signal removal through L2-regularization [32] using Matlab (R2013a, MathWorks, MA, USA), Bash (v4.2.25, Free Software Foundation, Boston, MA, USA) and MINC Toolkit (MINC tools, v2.0, McGonnell Brain Imaging Center, Montreal, QC, Canada). The resulting voxel spectra were quantified using LCModel (v6.3–1, LCMODEL Inc, ONT, CA) with a basis set of 14 components and a macromolecular baseline [33] in a spectral range of 1.8 - 4.2 ppm [15]. Due to their relevance in epilepsy, N-acetyl-aspartate and N-acetyl-aspartyl glutamate (tNAA), creatine and phosphocreatine (tCr), choline (tCho), myo-inositol (mIns), glutamate (Glu) and glutamine (Gln) were used in our analysis.

### 2.4. Data evaluation

Voxel-wise spectral quality was automatically assessed using the pseudo-replica method for SNR and full-width-at-half-maximum (FWHM) of tCr at 3.02 ppm and the fitting quality using the Cramér-Rao lower bounds (CRLBs) in all metabolites. Voxels were excluded if the tCr SNR < 5 or the tCr FWHM > 0.15 ppm [15].

In addition to the automized filtering of spectra, the overall quality of the metabolic maps was visually reviewed by a trained reader (S.C.) and referred to an MRI physicist (G.H.) in the presence of lipid- or movement artifacts. Metabolite maps that did not fulfill the quality criteria were excluded from further analysis. Equally, maps in which the EZ was located temporo-mesially were not included due to the interference of basal field inhomogeneities on our 7T scanner (**Figure 1**).

The remaining metabolite maps were then analyzed as ratio maps normalized to tNAA, because of three main advantages: due to the frequently described decrease of NAA in epilepsy, normalizing to this metabolite increases the detectability of potential hotspots; and second, as NAA is exclusively synthesized in neurons, normalization to NAA allows for a proxy for neuronal density even in pathological tissue with increased gliosis or atrophy. Last, in contrast to creatine, this metabolite has, to date, not been shown to be sensitive to clinical parameters such as seizure frequency. However, normalization to creatine does have the advantage of being relatively stable throughout the brain with a robust signal, for which reason a comparison of both ratios will be provided in this work.

To assess the sensitivity of MRSI in identifying altered metabolic status, metabolite maps were visually assessed in the region of interest (ROI), which represents the suspected EZ, as defined in the extensive clinical work-up compared to the contralateral hemisphere. Changes were categorically defined as the following:

- “increase”
- “decrease”
- “MRSI-negative” = no visible changes over the entirety of the assessed metabolite panel
- “stable” = no change in the EZ of a specific metabolite, despite alterations of other metabolites in the EZ

Metabolites of particular interest to our analysis were the ratios Glu, Gln, mIns, tCho, and tCr or NAA normalized to tNAA and tCr. In a second step, the metabolic patterns were qualitatively assessed in relation to the underlying pathologies and related to seizure frequency. Due to the role of Glu and Gln in energy metabolism, we further assessed their relationship in the epileptogenic zone.

## 3. Results

### 3.1. Detection of metabolic alterations in the EZ

Following the voxel-wise quality thresholding and visual assessment of the maps of the 53 patients who had completed the MRSI protocol (see **Figure 1**), 11 maps had to be excluded due to motion or lipid artifacts in the ROI and 13 due to basal locations affected by field inhomogeneities. The remaining 29 maps were comprised of 15 lesional and 14 MRI-negative cases. In the lesional group, metabolic alterations correlating to the suspected EZ were found in 86.67% of the maps in ratios normalized to NAA, whereas this was reduced to 80% when normalized to creatine (as illustrated in **Figure 2**). The assessed pathologies included eight cases of FCD, two mild malformations of cortical development (mMCD), two cases of polymicrogyria, one of mild malformation of cortical development with oligodendroglial hyperplasia and epilepsy (MOGHE), a patient with blurred GM/WM boundaries and chronic epilepsy associated changes and a patient with distinct focal atrophy of the temporolateral lobe with suspected MCD, a summary of metabolite trends is provided in **Table 2**. Histopathological verification of the above-mentioned pathologies was available in 66.67% (**Table 1**). In cases without histopathological assessment, the suspected MRI diagnosis was used as a classifier. The lesions that remained unidentified using MRSI included histopathologically confirmed MCD and suspected polymicrogyria. As illustrated in the exemplary spectra of a patient with FCD in **Figure 2**, metabolic patterns within the suspected EZ visibly differed from normal-appearing gray and white matter (NAGWM).

**Fig. 2.**
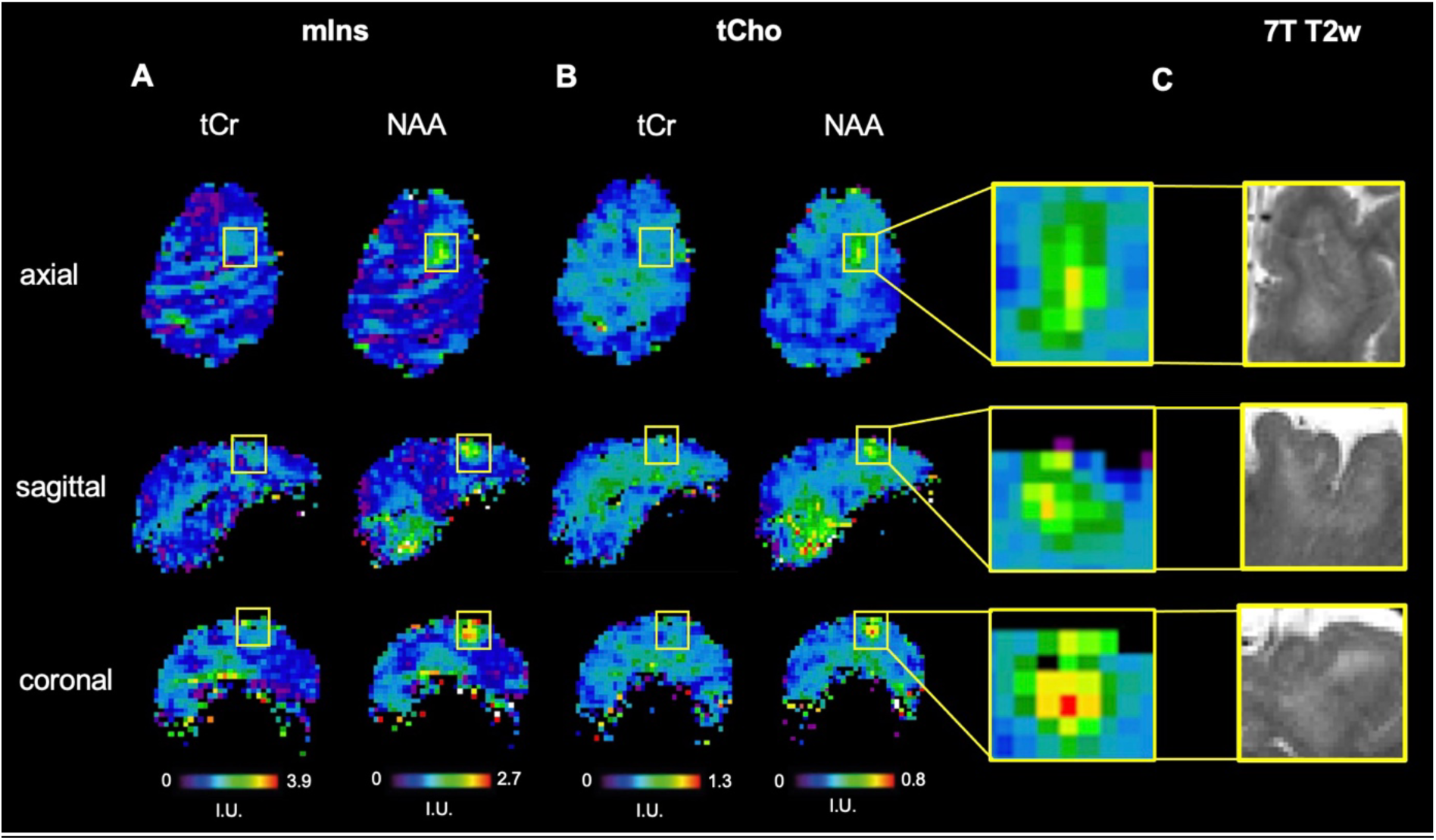
Ratio maps of mIns and tCho normalized to tCr and NAA showcasing the detectability of metabolic alterations in FCD. Metabolic ratio maps of patient 2 with FCD Type 2b. **A** mIns to tCr and tNAA and **B** tCho to tCr and tNAA. The corresponding lesions in 7T T2-weighted images are depicted in **C**. While the ratios normalized to NAA clearly show a metabolic hotspot corresponding to the lesion location, as highlighted in the yellow box, the structural abnormality remains nearly undetected in mIns/tCr and inconspicuous in tCho/tCr <u>Abbreviation</u>s: mIns = Myo-inositol, Cr = creatine, NAA = N-acetyl-aspartate + N-acetyl-aspartyl glutamate, tCho = total choline, 7T = 7 Tesla, T2w = T2-weighted

**Table 2:**
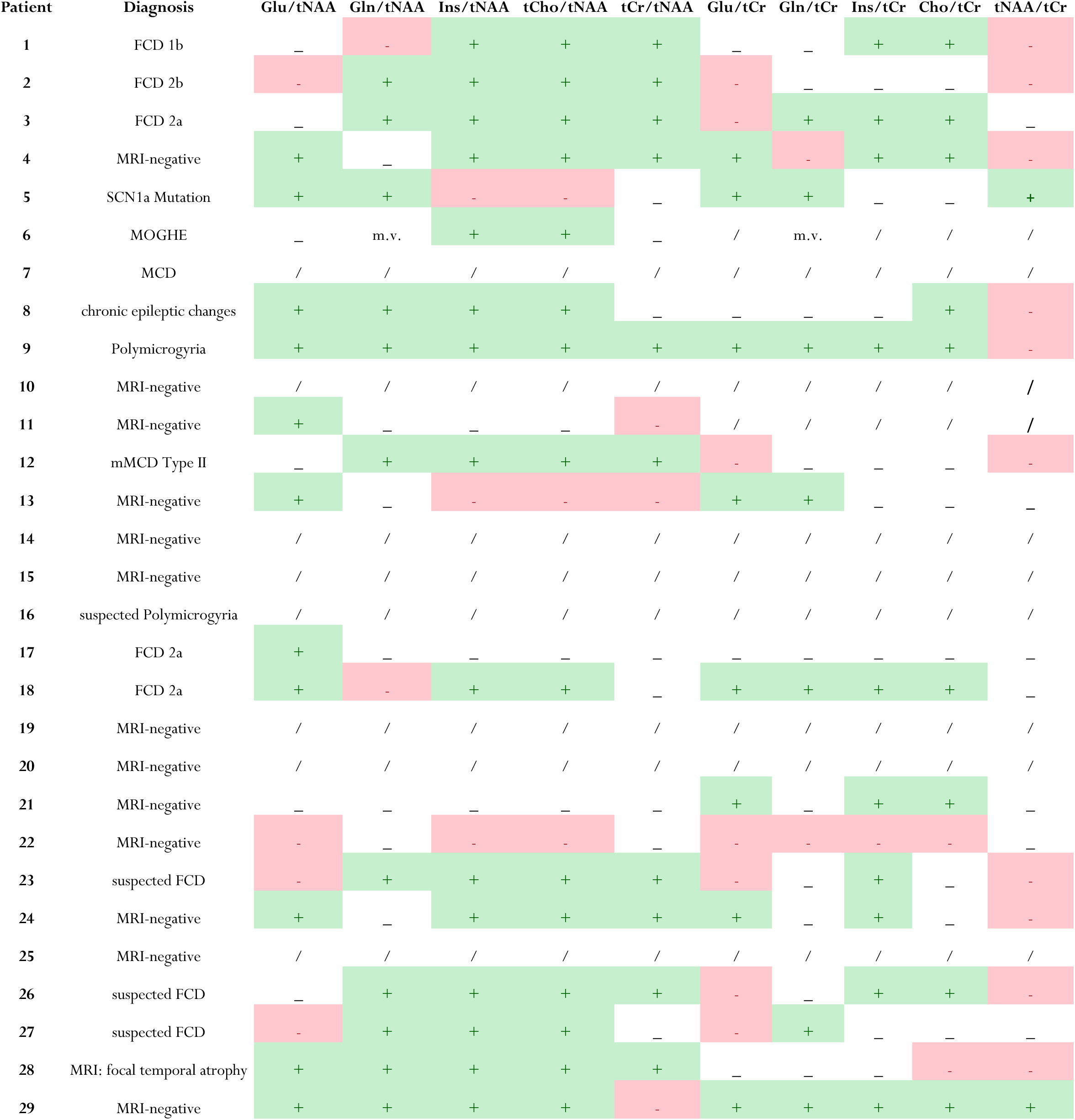
Summary of qualitative assessment of metabolite trends in EZ. + = increase in EZ, - = decrease in EZ, _= stable value of metabolites in EZ, / = MRSI-negative (no visual changes in all assessed metabolites in EZ), m.v. = missing value. Diagnosis refers to either histopathological assessment or MRI diagnosis (i.e., suspected). <u>Abbreviations:</u> mIns = Myo-inositol, Cr = creatine, NAA = N-acetyl-aspartate + N-acetyl-aspartyl glutamate, tCho = total choline, Glu = glutamate, Gln = glutamine, EZ = epileptogenic zone

Of the 14 PWE who were MRI-negative, 42.86% remained MRSI-negative. This highlights that measurable metabolic alterations were present in the majority of MRI-negative patients, although less circumscribed than in the presence of a visible lesion (**Suppl. Table 3**).

### 3.2. Stability of metabolite trends across pathologies

In a more detailed analysis of metabolite trends, we identified alterations over numerous pathologies (**Figure 3**), although no patterns were discernible due to the heterogeneity of the cohort and limited group sizes. However, within these changes we were able to distinguish a varying stability of metabolites, dependent on clinical parameters.

**Fig. 3.**
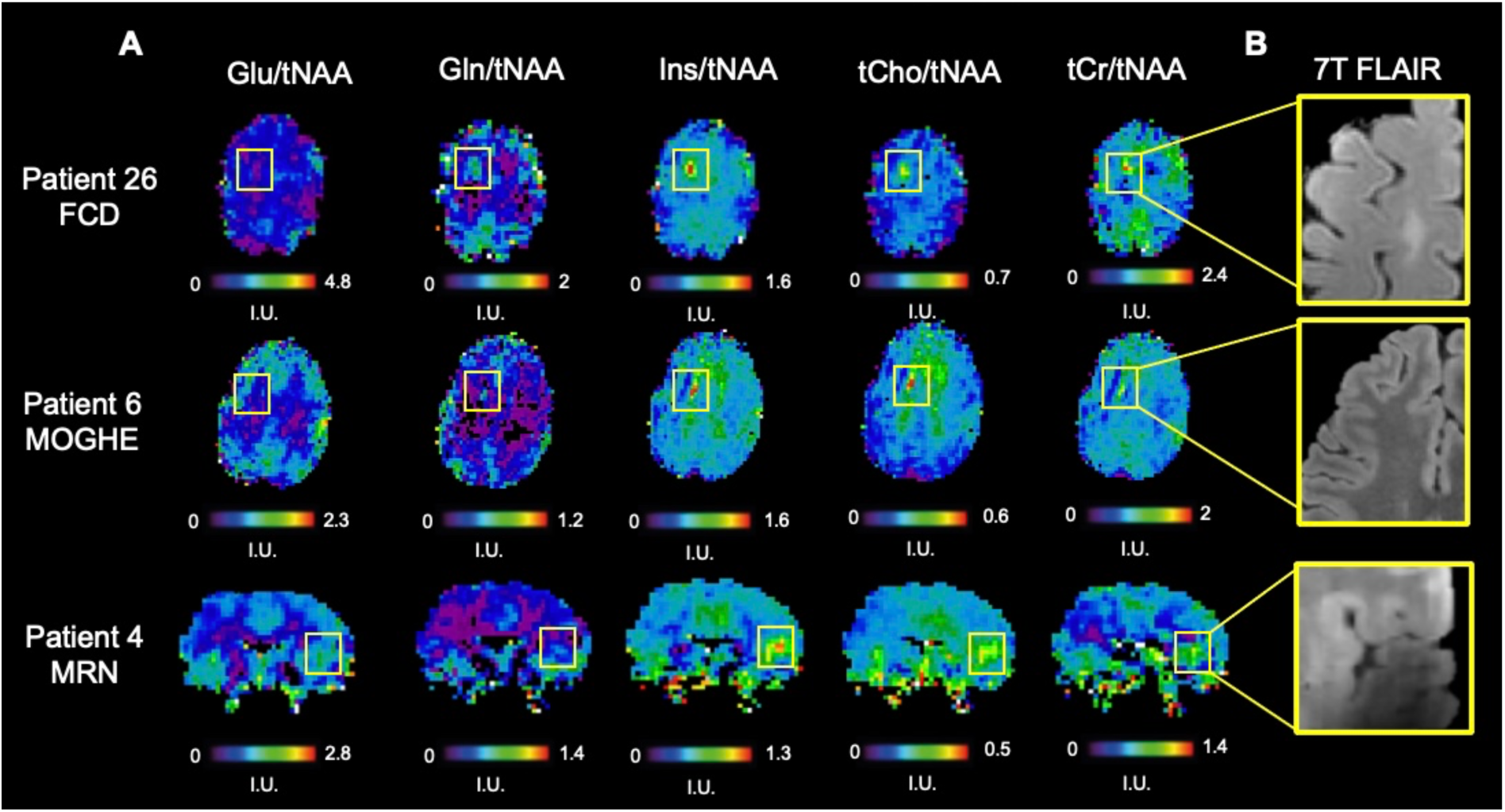
Overview of metabolic alterations in focal epilepsy. **A** Metabolic alterations are highlighted by the yellow box and the corresponding structural T2-weighted images are provided in **B**. <u>First row:</u> Axial metabolic ratio maps of patient 26 of Glu, Gln, Ins, tCho and tCr normalized to NAA with histopathologically verified FCD 2a. <u>Second row</u>: Axial metabolite ratio maps in patient 6 with MOGHE in the left frontal lobe. <u>Third row:</u> Coronal metabolite ratio maps in patient 4 with MRI-negative focal epilepsy. Based on the extensive clinical work-up, the suspected EZ was located in the right insula, in which MRSI showed metabolite ratios of mIns and tCho, albeit less circumscribed than in the lesionally confirmed patients. Field inhomogeneities visible in 7T FLAIR. <u>Abbreviations:</u> mIns = Myo-inositol, Cr = creatine, NAA = N-acetyl-aspartate + N-acetyl-aspartyl glutamate, tCho = total choline, 7T = 7 Tesla, T2w = T2-weighted, FCD = Focal Cortical Dysplasia, NAGWM = Normal appearing gray-and-white matter, MOGHE = Mild malformation of cortical development with oligodendroglial hyperplasia and epilepsy, MRN = MRI-negative, EZ = epileptogenic zone

Due to the overall higher detectability of alterations, further analysis was conducted only in maps normalized to NAA. In patients with visibly detectible lesions, circumscribed increases of tCho and mIns were observed in nearly all patients (92.31%) (excluding “MRSI negative” patients, i.e., those in whom none of the investigated metabolite ratios showed alterations), while no decreases in the suspected EZ were detected, as illustrated in **Figure 4**. Other fitted metabolites, such as Glu and Gln, showed varying directionality of changes, with Glu increases in 38.46%, decreases in 23.08%, and no changes in 38.46% of the investigated maps. Gln showed higher rates of increases (69.23%) and lower rates of decreases (15.38%). The proportion of maps with no visible alterations for Gln was 7.7%. Ratio maps of tCr/tNAA showed an increase in 61.54%, whereas in 38.46%, no changes were visually discernable.

**Fig. 4.**
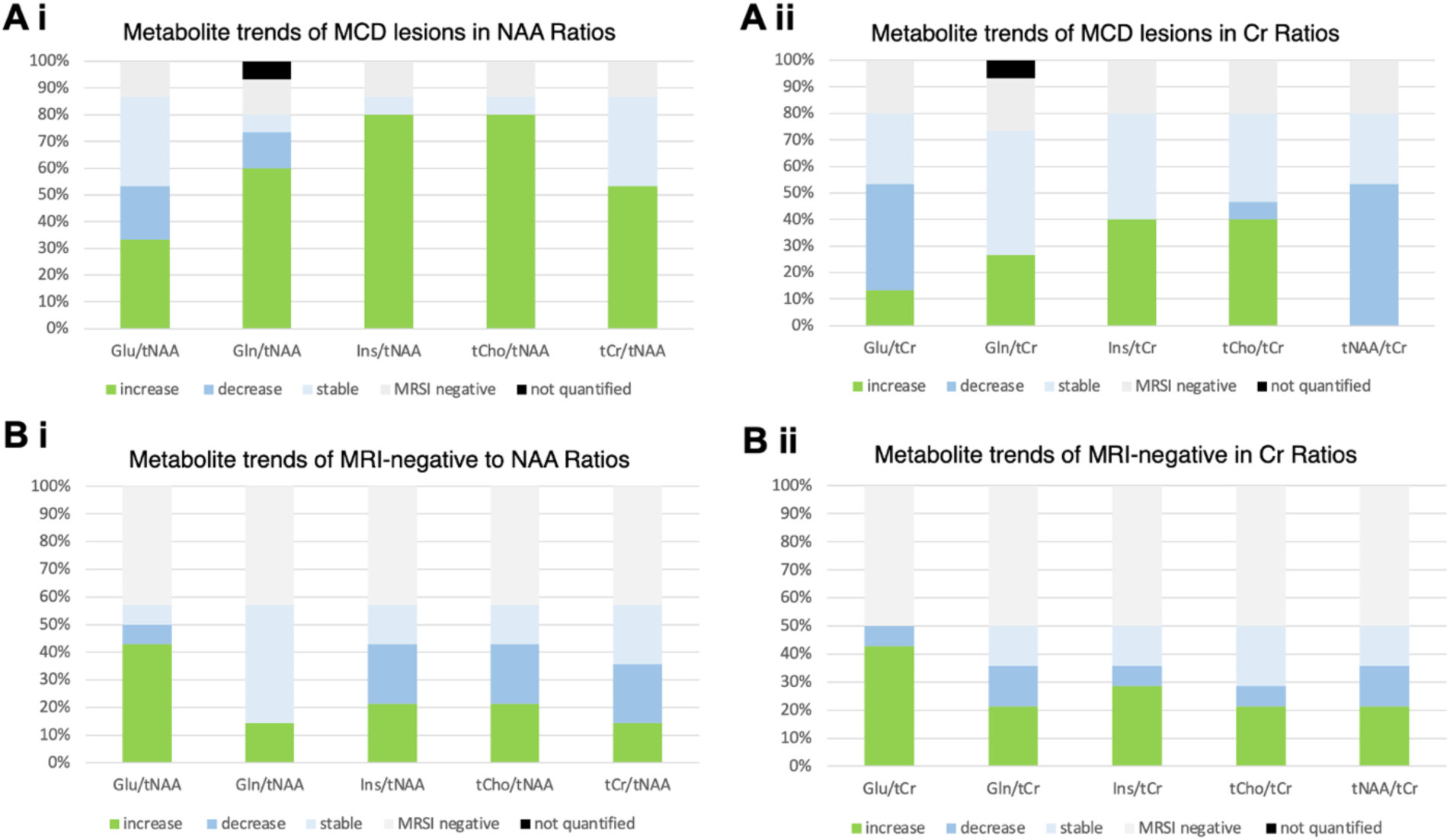
Metabolite trends in MCD lesions and MRI-negative cases. **Ai** and **Aii** illustrate the metabolite trends of Glu, Gln, mIns, tCho and tCr normalized to NAA and tCr respectively, showcasing the increased detection rate of metabolic alterations in ratios normalized to NAA. Specifically, normalizing to mIns and tCho show high sensitivity and stability in uncovering and characterizing MCD lesions. **Bi** and **Bii** in MRI-negative patients. In normalization to either metabolite, 60% showed alterations, albeit with heterogeneous patterns. Overall, normalizing to creatine uncovered markedly fewer metabolic alterations compared to NAA. “Stable” refers to no apparent change in the EZ of a specific metabolite, despite alterations of other metabolites in the ROI, whereas “MRSI-negative” refers to none of the investigated metabolites showing altered concentrations. <u>Abbreviations:</u> Glu = glutamate, Gln = glutamine, Ins = Myo-inositol, tCr = creatine, tNAA = N-acetyl-aspartate and N-acetyl-aspartyl glutamate, tCho = choline, ROI = region of interest, MCD = malformations of cortical development

The metabolic pattern in MRI-negative patients was markedly more heterogeneous, with an overall detection rate of 57.14%. Furthermore, all metabolites showed multi-directionality of changes (**Figure 4, B i and ii**), in contrast to the findings above. As histopathology and consequent postoperative seizure freedom of the suspected EZ was available in only one patient, the interpretation and implication of these results remain questionable.

Overall, metabolic maps showed a high heterogeneity of metabolic alterations throughout patients, summarized in **Suppl. Tables 2 and 3**. Furthermore, the trend of metabolite changes seems to be, at least partially, dependent on clinical parameters.

### 3.3. The relationship of glutamate to glutamine in the EZ

We next assessed the ratios of Glu/tNAA and Gln/tNAA in the epileptogenic zone compared to the contralateral hemisphere in relation to the seizure frequency; exemplary spectra are provided in **Figure 5**.

**Fig. 5.**
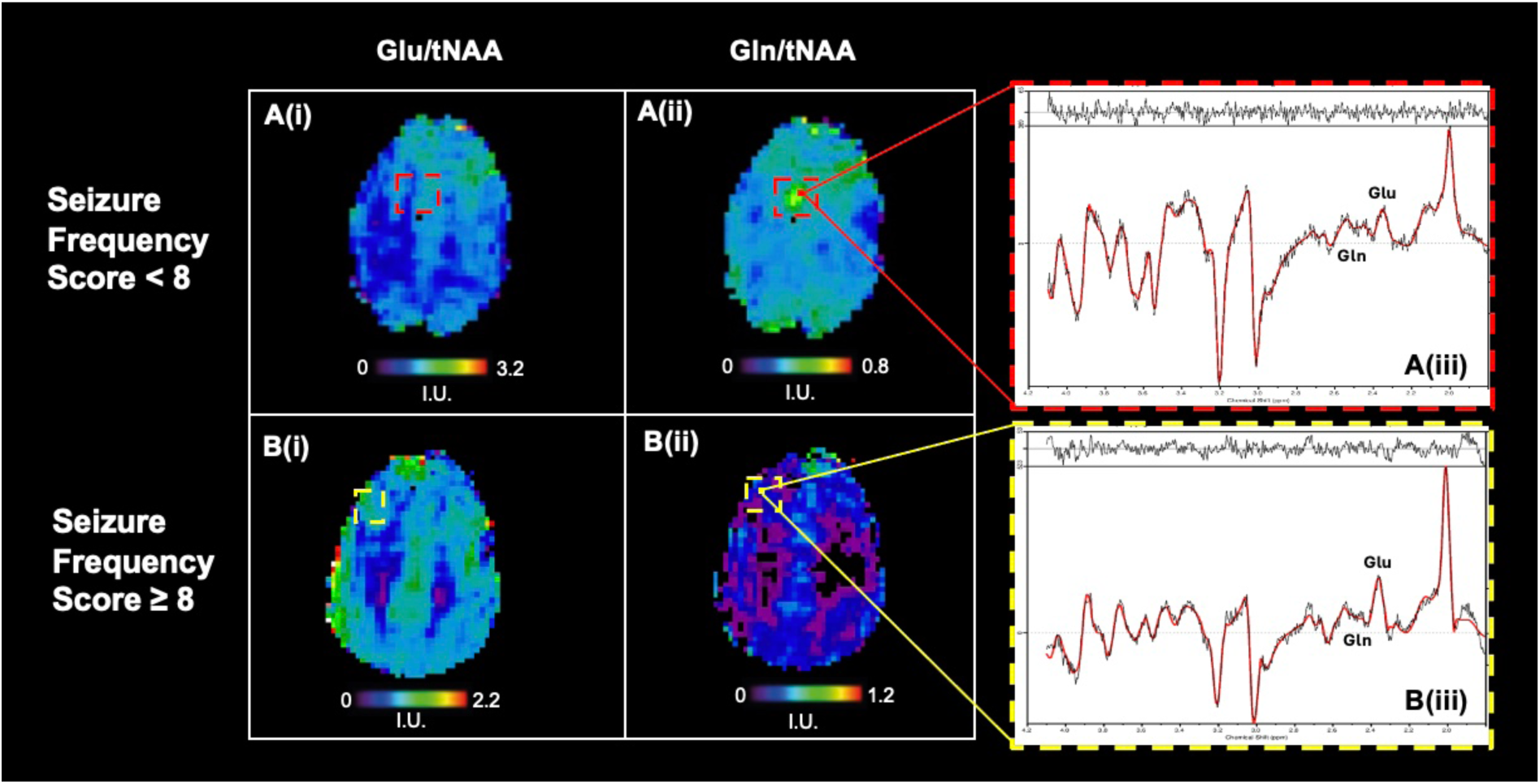
Glutamate and glutamine in relation to seizure frequency. Ratio maps of **(i)** Glu/tNAA, **(ii)** Gln/tNAA and **(iii)** corresponding sample spectra in the suspected EZ. **A:** Ratio maps of patient 23 with FCD in the left frontal lobe and SFS of 7 (i.e., monthly-occurring seizures) showing a relative decrease of glutamate and increase of glutamine in the ROI. **B:** Ratio maps of patient 18 with FCD 2a and SFS of 10 (i.e., daily occurring seizures). The sample spectra to the left showcase the subtle, although noticeable, relative increase of glutamate over glutamine when compared to **A(iii)**.

Specifically, we were able to observe that PWFE with lower seizure frequencies showed relative increases of glutamine and decreases of glutamate in the EZ, whereas a trend for the inverse was observable in patients with high seizure frequencies (i.e., weekly-occurring seizures and more, corresponding to SFS > 8), in which relative glutamate increases were more frequently measured. We observed glutamine increases in 71.43% of PWFE with SFS ≤ 8 (i.e., monthly-occurring seizures and less), whereas glutamate increases were observed only in 35.71%. Conversely, patients suffering from a high seizure burden more frequently showed glutamate increases (in 83.34%), in contrast to that of glutamine (in 16.67%). These trends are depicted in **Figure 6** and key findings are summarized in **Table 3**. Statistical analysis was conducted using Fisher’s Exact test. An extensive table providing the observed changes on an individual level in relation to the seizure frequency is provided in **Suppl. Table 4.**

**Fig. 6.**
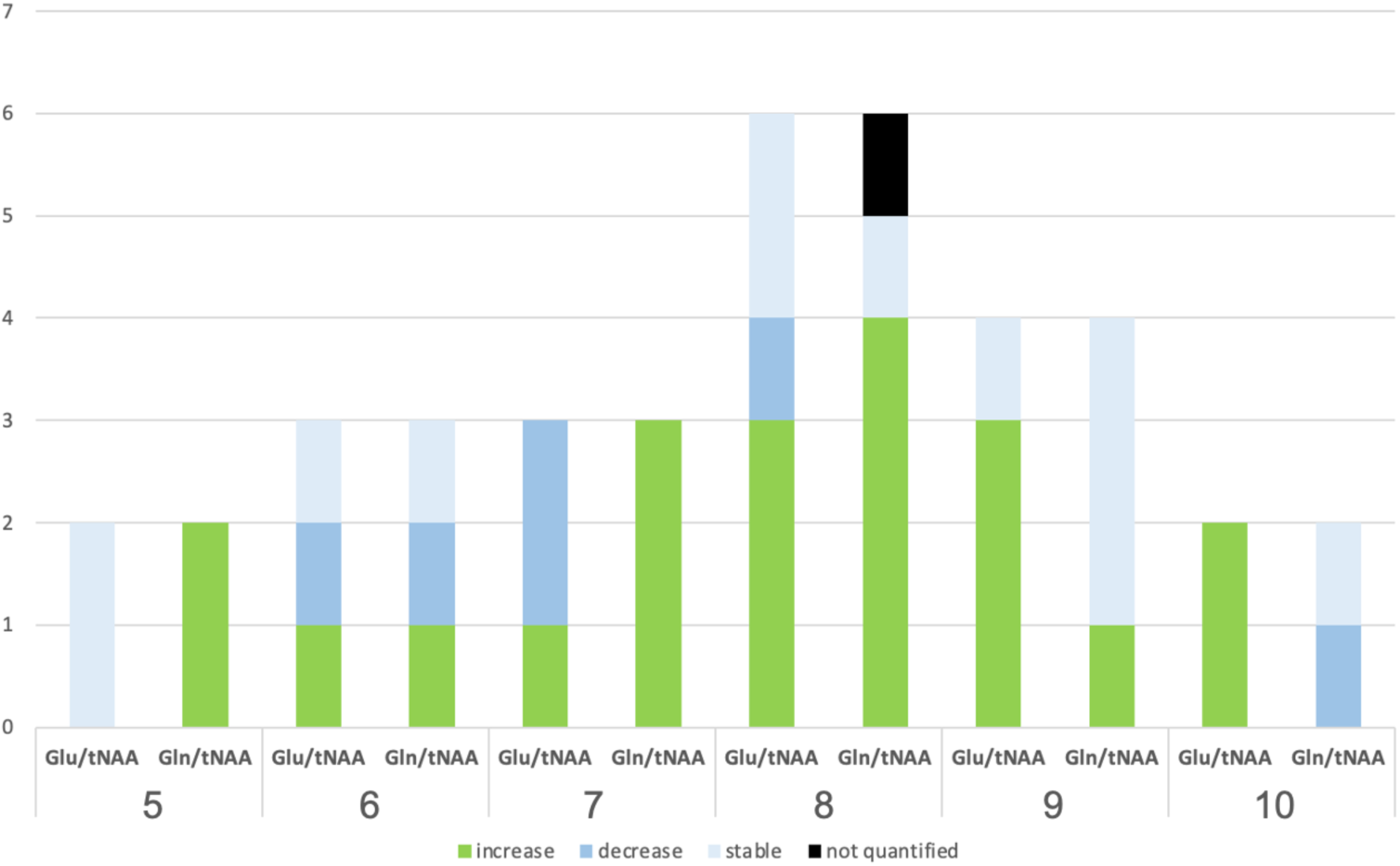
Trends of Glu and Gln in relation to the seizure frequency. x-axis: SFS ranging from 5 (yearly-occurring seizures) to 10 (daily-occurring seizures) y-axis: number of patients per group Preliminary visual inspection of the maps indicated a trend for Glu increases with higher seizure frequencies (i.e., daily-occurring seizures), whereas a tendency to the inverse, namely, Gln increases, were observed in PWFE with lower seizure frequencies (i.e., monthly-occurring seizures). <u>Abbreviations</u>: Glu = glutamate, Gln = glutamine, SFS = Seizure Frequency

**Table 3:** Summary of Key Findings. Top Row: Metabolite trends in PWFE with visibly detectable lesions excluding MRSI-negative cases. Bottom Row: Glu and Gln ratios in relation to the seizure frequency over the whole cohort. SFS < 8 signifies monthly-occurring seizures or less, whereas ≥ 8 corresponds to at least weekly-occurring seizures. One missing value for Gln in SFS < 8. p-values derived using Fisher’s Exact Test. <u>Abbreviations:</u> SFS = Seizure Frequency Score, mIns = myo-inositol, Cr = creatine, tNAA = N-acetyl-aspartate + N-acetyl-aspartyl glutamate, tCho = total choline, Glu = glutamate, Gln = glutamine

| Metabolite Trends |  |  |  |
| --- | --- | --- | --- |
|  | Increase | Decrease | Stable |
| Glu/tNAA | 38,46% | 23,08% | 38,46% |
| Gln/tNAA | 69,23% | 15,38% | 7,69% |
| tCh/tNAA | 92,31% | 0,00% | 7,69% |
| mIns/tNAA | 92,31% | 0,00% | 7,69% |
| tNAA/tCr | 61,54% | 0,00% | 38,46% |

**Table 3: Summary of Key Findings.**
| Relation to Seizure Frequency |  |  |  | p-value |
| --- | --- | --- | --- | --- |
| **SFS ≤ 8** | **Increase** | **Decrease** | **Stable** | 0,0572 |
| Glu/tNAA | 35,71% | 28,57% | 35,71% |  |
| Gln/tNAA | 71,43% | 7,14% | 4,17% |  |
| **SFS > 8** |  |  |  | 0,0801 |
| Glu/tNAA | 83,33% | 0,00% | 16,67% |  |
| Gln/tNAA | 16,67% | 16,67% | 66,67% |  |

## 4. Discussion

To the best of our knowledge, this is the first application of whole-brain ultra-high field MRSI at 7T in focal epilepsy. Although our preliminary qualitative analysis suggests that profiling of metabolic changes across pathologies remains challenging, 7T CRT-FID-MRSI shows promise in identifying metabolic alterations. Ratios normalized to NAA yielded an overall detection rate of 86.67% in the EZ, whereas this was reduced to 80% when normalized to tCr. This highlights that the NAA decrease is the main driver of the detection rate of metabolic alterations in our cohort. Of the assessed metabolites, the ratios mIns/tNAA and tCho/tNAA revealed the most metabolic alterations, whereas NAA/tCr uncovered changes in only 41.4%. In addition to assessing the acuity of MRSI in detecting metabolic alterations in focal epilepsy, this work provides some insight into the stability of these over pathologies and clinical parameters. We could observe that the directionality of change in assessed metabolites is at least partially dependent on clinical parameters such as the seizure frequency, most notably in glutamate and glutamine. Although previous work has also been able to demonstrate this for Cr [8], visual assessment of the maps was insufficient to uncover such changes or reach the level of statistical significance and will therefore require quantitative analysis. Conversely, metabolites that showed stable trends across pathologies included mIns, tCho and NAA. The discrepancy in influential factors of metabolic alterations is reflective of the nature of the assessed metabolites. Cr, Glu and Gln are inherently linked to energy and neurotransmitter metabolism [34], [35], whereas mIns and tCho are considered markers of structural remodeling. Contrasting this principle, NAA, although linked to energy metabolism, consistently shows a reduction in epilepsy, particularly in FDG-PET hypometabolic areas [17]. This may be indicative of a common denominator across some focal epilepsies, namely mitochondrial dysfunction. Recent research has shown dysfunctionality of the malate-aspartate shuttle [36] and impaired oxidative glucose metabolism in epilepsy [37], leading to a reduction of aspartate, the precursor to NAA. Neurons deprived of functional glucose metabolism may instead uphold energy metabolism by using glutamate as an alternative substrate for the tricarboxylic acid (TCA) cycle [25], a state which may be observed interictally. As neurons are unable to synthesize glutamate from glucose, *de novo* glutamate is instead synthesized in astrocytes and shuttled to neurons as glutamine, to be re-converted to glutamate and ready to be used as fuel in the TCA cycle [25]. Therefore, it may be plausible that, in neurons that hypometabolize glucose, a reduction of NAA, an increase of Gln, and a decrease of Glu could be expected in inter-ictal states in a compensatory attempt to uphold sufficient energy generation. Experimental rat models have demonstrated decreased NAA and increased Gln subsequent to status epilepticus [38] in inter-ictal states. This hypothesis is further supported by research investigating plasma concentrations of Glu and Gln, which has shown increased Glu plasma concentration levels in uncontrolled epilepsy, in contrast to elevated Gln plasma concentration levels when seizures are controlled [39]. Although all PWFE included in our study were drug-resistant, the seizure frequency included in our study spans from yearly- to daily-occurring seizures, allowing for a comparison within the cohort. Analogous to the findings above, we saw a relative increase of Glu to Gln in PWFE with a high seizure frequency, whereas relative Gln increases were observed in patients with low seizure frequencies in our preliminary analysis. An underlying confounder of this observation in our cohort may be the inter-ictal interval. Patients with high seizure frequencies were more likely to have had a seizure prior to the measurement and therefore potentially increased Glu levels as opposed to those with low seizure frequencies. Unfortunately, the timepoint of last seizure prior to measurement, necessary to validate this hypothesis, was not available during the course of this study. Further underpinning the complexity of energy metabolism in epilepsy, other groups have observed a slowing of the glutamate-glutamine cycle in hippocampi in mesial temporal epilepsy (17, 25), potentially contributing to delayed clearance of glutamate, and therefore higher glutamate concentrations following seizures. Nonetheless, characterization of glutamate and glutamine levels in epilepsy has thus far delivered divergent results as presented in a recent review [24] highlighting the necessity of interpreting these data in light of clinical measures, such as the timepoint of last seizure prior to measurement and seizure frequency.

### Limitations

Due to the limited cohort size inter-group comparisons across pathologies were not possible and larger cohort studies will enable a more detailed analysis of metabolic changes over various etiologies. Based on our preliminary results, the assessment of FDG-PET hypometabolism in relation to NAA and Gln levels may lend further insight into disease mechanisms. Furthermore, the validation of the correct clinical hypothesis of the epileptogenic zone in MRI-negative focal epilepsy is largely dependent on post-operative seizure freedom and, although this also holds true for lesional epilepsy, analysis of structural and functional changes is facilitated by the presence of a structural pathology. As only two MRI-negative patients were subsequently operated on, validation of the EZ hypothesis is limited, and therefore, also the assessed changes in the ROI as well. Moreover, the lack of knowledge of the last seizure prior to measurement impedes on our ability to interpret the observed changes. Energy metabolism is significantly affected by seizures and our results suggest that metabolic alterations are at least partially dependent on such clinical parameters. Furthermore, the effect of antiseizure medication was not considered in this analysis. However, as all PWFE included in this study were drug-resistant, most have had multiple therapeutic schemes, thereby potentially spreading the effect-size across the cohort. Future MRSI research, particularly of mono-therapeutic schemes, may lend insight into the metabolic effects of ASM. Last, the lack of comparable studies in this field limits the validity of our findings and future research may aid in contextualizing our results.

Methodologically, our analysis is limited by its qualitative nature and future work will focus on quantitative aspects, and therefore, the objective assessment of average metabolic alterations in segmented epileptogenic zones. Furthermore, B_0_-inhomogeneities, inherent to the ultra-high-field MRSI research, severely impeded the assessment of basal locations, such as mesial temporal lobes, essential in epilepsy research, necessitating their exclusion from further analysis within the scope of this study.

## Conclusion

In this study, we could successfully generate high-resolution, whole-brain maps in epilepsy using 7T CRT-FID MRSI. Furthermore, we could show that this technique yields high detection rates of metabolic alterations across multiple pathologies corresponding to the EZ and lend some insight into the stability of metabolic alterations in relation to clinical parameters. Although we were not able to demonstrate group differences across etiologies, our findings suggest that, while metabolites such as mIns and tCho show stable trends of changes, the directionality of alterations in Glu and Gln show a dependency on the seizure frequency. Furthermore, we propose that Glu and Gln share an inverse relationship in the EZ dependent on the seizure interval, highlighting the necessity of spectral separation of these metabolites. Quantitative analysis will be necessary for more robust statistical representation in future work. With the availability of new MRSI methods at 7T, *in vivo* validation of this hypothesis is possible for the first time and further work in this area by multiple groups may greatly contribute to understanding disease mechanisms. Conclusively, our preliminary analysis suggests that 7T CRT-FID MRSI shows promise not only as a feasible method for detecting metabolic alterations, but also may further aid in understanding underlying complex metabolic pathomechanisms in epilepsy.

## Data Availability

All data produced in the present work are contained in the manuscript

## Acknowledgments

This research was funded in whole, or in part, by the Austrian Science Fund (FWF) grant 10.55776/KLI1121. For open access purposes, the author has applied a CC BY public copyright license to any author-accepted manuscript version arising from this submission. We were further supported by the Medical Scientific Fund of the Mayor of the Federal Capital Vienna (Project Number 21186). The financial support by the Austrian Federal Ministry for Digital and Economic Affairs and the National Foundation for Research, Technology and Development, and the Christian Doppler Research Association, is gratefully acknowledged.

## Abbreviations

tCho: choline-containing compounds
tCr: total creatine, creatine + phosphocreatine
CRLB: Cramér-Rao lower bound
CRT: concentric ring trajectories
EEG: electroencephalography
EZ: epileptogenic zone
FCD: focal cortical dysplasia
FID: free induction decay
FLAIR: fluid-attenuated inversion recovery
FOV: field of view
FWHM: full width at half maximum
Gln: glutamine
Glu: glutamate
HS: hippocampal sclerosis
LEAT: low-grade epilepsy-associated neuroepithelial tumour
MCD: malformations of cortical development
mIns: myo-inositol
M2RAGE: magnetization-prepared 2 rapid acquisition gradient echoes
MRI: magnetic resonance imaging
MRSI: magnetic resonance spectroscopic imaging
tNAA: N-acetyl-aspartate + N-acetyl-aspartyl glutamate
PET: positron emission tomography
PW(F)E: people with (focal) epilepsy
ROI: region of interest
SAR: specific absorption rate
SNR: signal-to-noise ratio
UHF: ultra-high-field
WET: water suppression enhanced through T_1_ effects
WMS: fluid and white-matter suppressed

## Statements and Declarations

The authors have no financial or non-financial interests to declare that are directly or indirectly related to this work.

