## Supplemental data for "The relationship of glutamate and glutamine and metabolic profiling in focal epilepsy using 7T CRT-FID-MRSI"

| Patient | Age | Sex | Epilepsy Duration | Seizure Frequency | Suspected EZ | MRI diagnosis | FDG-PET Hypometabolism | Histopathology |
|---|---|---|---|---|---|---|---|---|
| **1** | 21-25 | male | 20 | 6 | R frontal | suspected FCD R frontal | R frontal | FCD Ib |
| **2** | 31-35 | male | 17 | 8 | R frontal | suspected FCD R frontal | - | FCD IIb |
| **3** | 26-30 | female | 10 | 8 | R parietal | suspected FCD R centroparietal | R frontoparietal | FCD IIa |
| **4** | 21-25 | male | 17 | 9 | R frontal | MRI-negative | R insular and temporolateral | |
| **5** | 21-25 | female | 24 | 6 | R hemisphere | MRI-negative | negative | |
| **6** | 26-30 | female | 4 | 7 | L frontal | suspected FCD L frontal | L frontal | MOGHE |
| **7** | 16-20 | female | 12 | 7 | R temporal | suspected blurring R temporal | R temporal | Mild malformation |
| **8** | 21-25 | male | 19 | 9 | R temporal | signal alteration R frontal | R temporal | chronic epilepsy-associated changes, blurred G/WM border |
| **9** | 31-35 | female | 9 | 8 | L parietooccipital | migration disorder L parieto-occipital | L parieto-occipital | Polymicrogyria |
| **10** | 31-35 | male | 20 | 7 | Lhemisphere | MRI-negative | negative | |
| **11** | 16-20 | female | 9 | 2 | occipital | MRI-negative | negative | |
| **12** | 16-20 | male | 6 | 7 | R temporal | MRI-negative | negative | mMCD Typ II |
| **13** | 31-35 | male | 25 | 10 | L postcentral | MRI-negative | L insula | |
| **14** | 36-40 | male | 6 | 8 | R hemisphere | MRI-negative | R temporal | |
| **15** | 12-15 | male | 6 | 9 | L frontal | MRI-negative | negative | |
| **16** | 12-15 | female | 7 | 8 | bitemporal | suspected bilateral perisylvic polymicrogyria | L temporal | |
| **17** | 21-25 | female | 20 | 9 | R frontal | bilateral temporal signal alteration | R frontal | FCD IIa |
| **18** | 12-15 | female | 2 | 10 | L frontal | suspected bottom-of-sulcus dysplasia L frontal | negative | FCD IIb |
| **19** | 26-30 | female | 8 | 9 | multiregional | MRI-negative | negative | |
| **20** | 31-35 | male | 18 | 8 | frontal | MRI-negative | negative | |
| **21** | 31-35 | female | 10 | 6 | L temporal | MRI-negative | negative | |
| **22** | 21-25 | male | 12 | 8 | R temporo-parietal | MRI-negative | negative | |
| **23** | 26-30 | male | 18 | 7 | L frontal | signal alteration L frontal | L frontal | |

| | | | | | | | |
|---|---|---|---|---|---|---|---|
| **24** | 16-20 | male | 12 | 8 | temporal | MRI-negative | negative |
| **25** | 56-60 | female | 27 | 7 | R frontal | MRI-negative | R temporal |
| **26** | 12-15 | male | 9 | 5 | L frontal | suspected FCD L frontal | L frontal |
| **27** | 16-20 | female | 15 | 7 | R frontal | suspected FCD R frontal | R frontolateral |
| **28** | 31-35 | male | 19 | 8 | R temporolateral | focal atrophy R temporolateral | R temporolateral |
| **29** | 41-45 | male | 25 | 7 | R temporal plus | MRI-negative | L temporal |

Supplementary Table 1: **Clinical data of included patients.** All patients underwent prolonged video-EEG-monitoring at a tertiary epilepsy center for the assessment of the suspected EZ. This also included the assessment of the seizure semiology, clinical 3T MRI with a dedicated epilepsy protocol, neuropsychological testing and FDG-PET. The seizure frequency was estimated using the Seizure Frequency Score [29]. MRI-negative excluded incidental findings unrelated to the suspected EZ. Where available, histopathological verification of the suspected pathology is supplied. In cases without histopathological assessment, the radiological diagnosis was used as a classifier.

Abbreviations: FCD = Focal Cortical Dysplasia, R = right, L = left, EZ = epileptogenic zone

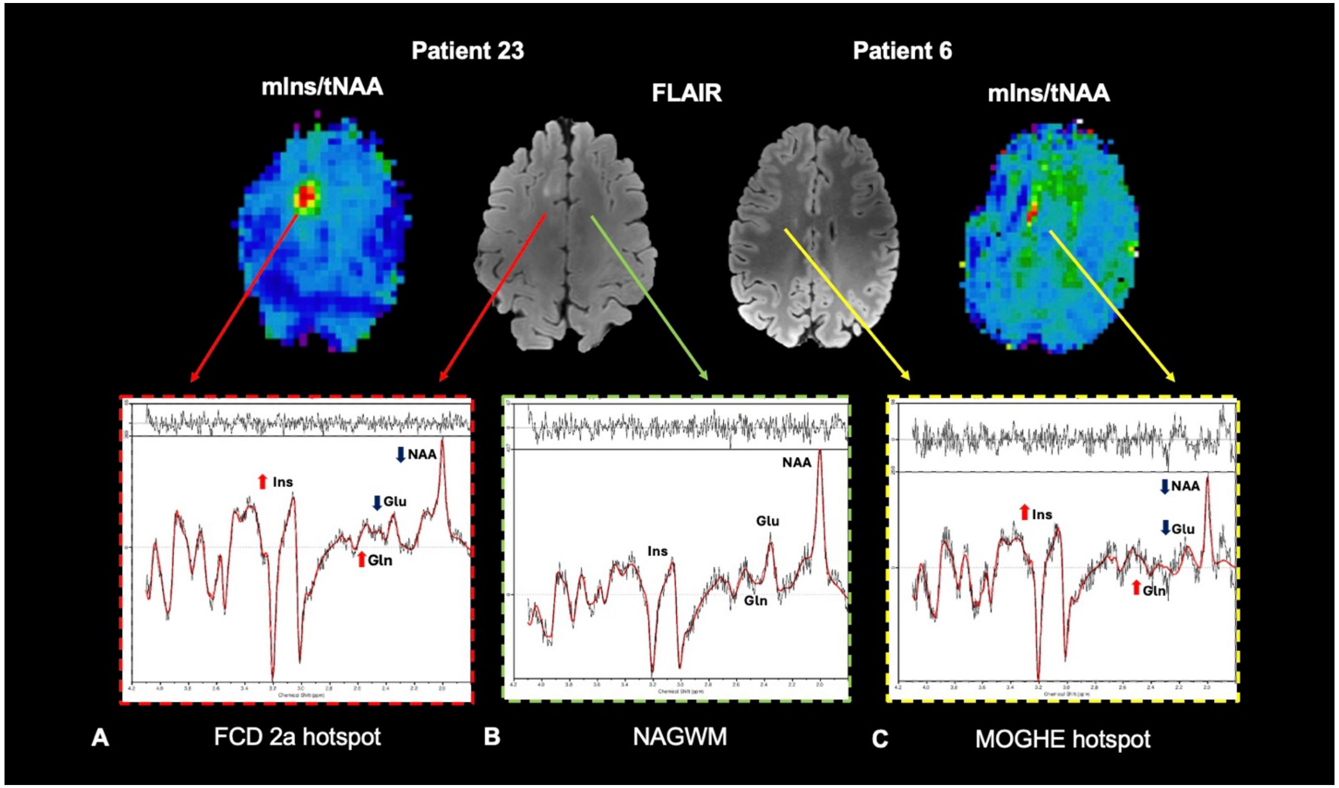


**Supplementary Fig.1 Exemplary spectra**
**A** histopathologically verified FCD
**B** normal-appearing gray-matter contralaterally and
**C** histopathologically verified MOGHE in the left frontal lobe. The red arrows point to the corresponding spectra of the FCD lesion, the green arrow to spectra in normal-appearing gray-matter, and the yellow arrows to exemplary spectra in an MOGHE hotspot. T2-weighted images of the lesions are provided in the center.
<u>Abbreviations:</u> FCD = Focal Cortical Dysplasia, NAGWM = Normal-appearing gray and white matter, MOGHE = Mild Malformation of Cortical Development with Oligodendroglial Hyperplasia and Epilepsy

| Patient | Histopathology | FDG-PET Hypometabolism | suspected EZ | SFS | Duration | Glu/tNAA | Gln/tNAA | Ins/tNAA | tCho/tNAA | tCr/tNAA |
|---|---|---|---|---|---|---|---|---|---|---|
| **1** | FCD 1b | R frontal | R frontal | 6 | 20 | _ | - | + | + | + |
| **2** | FCD 2b | - | R frontal | 8 | 17 | - | + | + | + | + |
| **3** | FCD 2a | R parietal | R parietal | 8 | 10 | _ | + | + | + | + |
| **6** | MOGHE | L frontal | L frontal | 8 | 4 | _ | m.v. | + | + | _ |
| **7** | MCD | R temporal | R temporal | 7 | 12 | / | / | / | / | / |
| **8** | chronic epileptic changes | R temporal | R temporal | 9 | 19 | + | + | + | + | _ |
| **9** | Polymicrogyria | L parietooccipital | L parietal | 8 | 9 | + | + | + | + | + |
| **12** | mMCD Type II | negative | R temporal | 5 | 6 | _ | + | + | + | + |
| **16** | suspected Polymicrogyria | L mesiotemporal | bitemporal | 8 | 7 | / | / | / | / | / |
| **17** | FCD 2a | R frontal | R frontal | 9 | 20 | + | _ | _ | _ | _ |
| **18** | FCD 2a | negative | L frontal | 10 | 2 | + | - | + | + | _ |
| **23** | suspected FCD | L frontal | L frontal | 7 | 18 | - | + | + | + | + |
| **26** | suspected FCD | L frontal | L frontal | 5 | 9 | _ | + | + | + | + |
| **27** | suspected FCD | R frontal | R frontal | 7 | 15 | - | + | + | + | _ |
| **28** | MRI: focal temporal atrophy | R temporolateral | R temporolateral | 8 | 19 | + | + | + | + | + |

Supplementary Table 2: **Metabolite trends in NAA ratios in lesional epilepsy.**
+ = increase in EZ, - = decrease in EZ, _= stable value of metabolite in EZ, / = MRSI-negative (no visual changes in all assessed metabolites in EZ), m.v. = missing value.
Abbreviations: R = right, L = left, mIns = Myo-inositol, Cr = creatine, NAA = N-acetyl-aspartate + N-acetyl-aspartyl glutamate, tCho = total choline, Glu = glutamate, Gln = glutamine, EZ = epileptogenic zone

| Patient | Histopathology | FDG-PET Hypometabolism | suspected EZ | SFS | Duration | Glu/tNAA | Gln/tNAA | Ins/tNAA | tCho/tNAA | tCr/tNAA |
|---|---|---|---|---|---|---|---|---|---|---|
| **4** | - | R insula | R frontal | 9 | 17 | + | _ | + | + | + |
| **5** | SCN1a Mutation | negative | R hemispheric | 6 | 24 | + | + | - | - | _ |
| **10** | - | negative | L hemispheric | 7 | 20 | / | / | / | / | / |
| **11** | - | negative | occipital | 2 | 9 | + | _ | _ | _ | - |
| **13** | - | L insula | undefined | 10 | 25 | + | _ | - | - | - |
| **14** | - | R temporal | R hemisphere | 8 | 6 | / | / | / | / | / |
| **15** | - | negative | L opercular | 9 | 6 | / | / | / | / | / |
| **19** | - | negative | multiregional | 9 | 8 | / | / | / | / | / |
| **20** | - | negative | L frontal | 8 | 18 | / | / | / | / | / |
| **21** | - | negative | L temporal | 9 | 10 | _ | _ | _ | _ | _ |
| **22** | - | negative | R temporoparietal | 6 | 4 | - | _ | - | - | _ |
| **24** | - | negative | R temporal/parietal | 8 | 12 | + | _ | + | + | + |
| **25** | - | R temporobasal | R frontal | 7 | 27 | / | / | / | / | / |
| **29** | - | L temporal | R temporal plus | 7 | 25 | + | + | + | + | - |

Supplementary Table 3: **Metabolite trends in NAA ratios in MRI-negative PWFE.**
+ = increase in EZ, - = decrease in EZ, _= stable value of metabolites in EZ, / = MRSI-negative (no visual changes in all assessed metabolites in EZ), m.v. = missing value.
Abbreviations: R = right, L = left, mIns = Myo-inositol, Cr = creatine, NAA = N-acetyl-aspartate + N-acetyl-aspartyl glutamate, tCho = total choline, Glu = glutamate, Gln = glutamine, EZ = epileptogenic zone

| Patient | Histopathology | FDG-PET Hypometabolism | suspected EZ | SFS | Duration | Glu/tNAA | Gln/tNAA | Ins/tNAA | tCho/tNAA | tCr/tNAA |
|---|---|---|---|---|---|---|---|---|---|---|
| **11** | - | negative | occipital | 2 | 9 | + | _ | _ | _ | - |
| **12** | mMCD Type II | negative | R temporal | 5 | 6 | _ | + | + | + | + |
| **26** | MRI: FCD | L frontal | L frontal | 5 | 9 | _ | + | + | + | + |
| **5** | SCN1a Mutation | negative | R hemispheric | 6 | 24 | + | + | - | - | _ |
| **22** | - | negative | R temporoparietal | 6 | 4 | - | _ | - | - | _ |
| **1** | FCD 1b | R frontal | R frontal | 6 | 20 | _ | - | + | + | + |
| **10** | - | negative | L hemispheric | 7 | 20 | / | / | / | / | / |
| **25** | - | R temporobasal | R frontal | 7 | 27 | / | / | / | / | / |
| **29** | - | L temporal | R temporal plus | 7 | 25 | + | + | + | + | - |
| **7** | MCD | R temporal | R temporal | 7 | 12 | / | / | / | / | / |
| **23** | MRI: FCD | L frontal | L frontal | 7 | 18 | - | + | + | + | + |
| **27** | MRI: FCD | R frontal | R frontal | 7 | 15 | - | + | + | + | _ |
| **14** | - | R temporal | R hemisphere | 8 | 6 | / | / | / | / | / |
| **20** | - | negative | L frontal | 8 | 18 | / | / | / | / | / |
| **24** | - | negative | R temporal/parietal | 8 | 12 | + | _ | + | + | + |
| **2** | FCD 2b | - | R frontal | 8 | 17 | - | + | + | + | + |
| **3** | FCD 2a | R parietal | R parietal | 8 | 10 | _ | + | + | + | + |
| **6** | MOGHE | L frontal | L frontal | 8 | 4 | _ | m.v. | + | + | _ |
| **9** | Polymicrogyria | L parietooccipital | L parietal | 8 | 9 | + | + | + | + | + |
| **16** | MRI: Polymicrogyria | L mesiotemporal | bitemporal | 8 | 7 | / | / | / | / | / |

| | | | | | | | | | | |
|---|---|---|---|---|---|---|---|---|---|---|
| **28** | MRI: focal temporal atrophy | R temporolateral | R temporolateral | 8 | 19 | + | + | + | + | + |
| **4** | - | R insula | R frontal | 9 | 17 | + | _ | + | + | + |
| **15** | - | negative | L opercular | 9 | 6 | / | / | / | / | / |
| **19** | - | negative | multiregional | 9 | 8 | / | / | / | / | / |
| **21** | - | negative | L temporal | 9 | 10 | _ | _ | _ | _ | _ |
| **8** | chronic epileptic changes | R temporal | R temporal | 9 | 19 | + | + | + | + | _ |
| **17** | FCD 2a | R frontal | R frontal | 9 | 20 | + | _ | _ | _ | _ |
| **13** | - | L insula | undefined | 10 | 25 | + | _ | - | - | - |
| **18** | FCD 2a | negative | L frontal | 10 | 2 | + | - | + | + | _ |

Supplementary Table 4: **Metabolite trends in NAA ratios with increasing seizure frequency**.

Symbols: + = increase in EZ, - = decrease in EZ, _= stable value of metabolites in EZ, / = MRSI-negative (no visual changes in all assessed metabolites in EZ), m.v. = missing value.

Abbreviations: R = right, L = left, mIns = Myo-inositol, Cr = creatine, NAA = N-acetyl-aspartate + N-acetyl-aspartyl glutamate, tCho = total choline, Glu = glutamate, Gln = glutamine, EZ = epileptogenic zone

| Minimum Reporting Standards in MR Spectroscopy Overview | |
|---|---|
| **Site** | Vienna High-Field MR Center |
| **1. Hardware** | |
| **a. Field strength** | 7T |
| **b. Manufacturer** | Siemens |
| **c. Model** | Magnetom |
| **d. RF coils: nuclei (transmit/ receive), number of channels, type, body part** | 1H, 32 ch, head, Nova Medical |
| **e. Additional hardware** | N/A |
| **2. Acquisition** | |
| **a. Pulse sequence** | FID-MRSI |
| **b. Volume of interest (VOI) locations** | lesion, EZ, NAGWM |
| **c. Nominal VOI size** | 220×220×110 mm³ |
| **d. Repetition time (TR), echo time (TE)** | 450 ms / 1.3 ms acquisition delay |
| **e. Total number of excitations or acquisitions per spectrum** | N/A, spatial-spectral encoding |
| **In-time series for kinetic studies** | N/A |
| **i. Number of averaged spectra (NA) per time-point** | N/A |
| **ii. Averaging method (e.g., block-wise or moving average)** | N/A |
| **iii. Total number of spectra (acquired / in-time series)** | N/A |
| **f. Additional sequence parameters (spectral width in Hz, number of spectral points, frequency offsets); If STEAM: Mixing Time TM; If MRSI: 2D or 3D, FOV in all directions, matrix size, acceleration factors, sampling method** | BW 2778 Hz, 1920 spectral points, MRSI: 3D, 220×220×133 mm³, 64×64×39, spatial-spectral encoding |
| **g. Water suppression method** | WET |
| **h. Shimming method, reference peak, and thresholds for "acceptance of shim" chosen** | Standard shim + manual adjustment, water peak < 50 Hz |
| **i. Triggering or motion correction method** | N/A |
| **3. Data analysis methods and outputs** | |
| **a. Analysis software** | LCModel 6.3-1 |
| **b. Processing steps deviating from quoted reference or product** | N/A |
| **c. Output measure** | Ratio |
| **d. Quantification references and assumptions, fitting model assumptions** | Simulated in NMRScope-B, macromolecular background |
| **4. Data Quality** | |
| **a. Reported variables (SNR, linewidth (with ref. peaks))** | SNR and linewidths not reported |
| **b. Data exclusion criteria** | tCr SNR <5; tCr FWHM >0.15 ppm |
| **c. Quality measures of post processing model fitting** | CRLB |
| **d. Sample spectrum** | See Supp. Fig. 1 |

Supplementary Table 4: Overview of Minimum Reporting Standards for *in vivo* Microscopy used in this study [30].
